# Hydroxychloroquine has no effect on SARS-CoV-2 load in nasopharynx of patients with mild form of COVID-19

**DOI:** 10.1101/2020.06.30.20143289

**Authors:** Alexey Komissarov, Ivan Molodtsov, Oxana Ivanova, Elena Maryukhnich, Svetlana Kudryavtseva, Alexey Mazus, Evgeniy Nikonov, Elena Vasilieva

**Author notes:** Corresponding author: Elena Vasilieva, MD, PhD, Clinical City Hospital named after I.V. Davydovsky, 11 Yauzskaya Street, Moscow, Russia, 109240.

## Abstract

Due to the constantly growing numbers of COVID-19 infections and death cases attempts were undertaken to find drugs with anti SARS-CoV-2 activity among ones already approved for other pathologies. In the framework of such attempts, in a number of in vitro, as well as in vivo, models it was shown that hydroxychloroquine (HCQ) has an effect against SARS-CoV-2. While there was not enough clinical data to support the use of HCQ, several countries including Russia have included HCQ in treatment protocols for infected patients and for prophylactic. Here, we evaluated the SARS-CoV-2 RNA in nasopharynx swabs from infected patients in mild conditions and compared the viral RNA load dynamics between patients receiving HCQ and control group without antiviral pharmacological therapy. We found statistically significant relationship between maximal RNA quantity and patients’ deteriorating medical conditions, as well as confirmed the arterial hypertension to be a risk factor for people with COVID-19. However, we showed that HCQ therapy neither shortened the viral shedding period nor reduced the virus RNA load.

## Introduction

Currently, the number of SARS-CoV-2 infection cases worldwide has exceeded 29 million including more than 930 thousand registered deaths. Due to the constantly growing numbers of infections and death cases it is paramount to find efficient antivirals that block the SARS CoV-2 infection. Since *de novo* drug development is a very long and complex process, attempts were undertaken to find drugs with anti SARS-CoV-2 activity among ones already approved for other pathologies. In the framework of such attempts, in a number of *in vitro*, as well as *in vivo*, models it was shown that a well-known and widely available compound hydroxychloroquine (HCQ) has an effect against SARS-CoV-2 [1-3]. Furthermore, it was reported on the use of HCQ for SARS-CoV-2 patients with severe disease, but the results of these studies are highly controversial. While there was not enough clinical data to support the use of HCQ, several countries including Russia have included HCQ in treatment protocols for infected patients and for prophylactic. Here, we evaluated the SARS-CoV-2 RNA in nasopharynx swabs from infected patients in mild conditions and compared the viral RNA load dynamics between patients receiving HCQ and control group without antiviral pharmacological therapy.

## Materials and Methods

A total of 45 patients with COVID-19 in mild condition were enrolled in the study. Among them 33 patients were receiving hydroxychloroquine (200 mg twice a day) while 12 patients represented a control group (receiving no antiviral pharmacological therapy). Local ethics committees approved the study protocol and all participants provided their written consent. The patients were included into the study 7-10 days after the onset of symptoms and if their nasopharynx swab was positive for SARS-CoV-2.

All patients were regularly examined by a doctor and the severity of symptoms was registered. Patient’s body temperature holding higher than 38 °C for 4 days or more and/or the blood oxygen saturation level dropping lower than 95% were considered as condition deteriorating. In this case the patient was either hospitalized or underwent intense home observation depending on the patient’s overall state. At first visit nasopharynx swab and peripheral blood were collected from each patient, then nasopharynx swabs were collected at day 3 and 8.

Peripheral blood collected from the forearm vein into S-Monovette 2.7 mL K3E and 4.9 mL Z tubes (Sarstedt, Germany) was analyzed for complete blood count and biochemical panel, respectively, using automated procedures.

After probing nasopharynx swabs were placed into viral transport media (COPAN Diagnostics, USA), transported at 4 °C then stored at −20 °C. Viral RNA was isolated using RIBO-prep kit (AmpliSens, Russia) according to the manufacturer protocol. Briefly, 100 μl of thawed transport media were lysed, then nucleic acids were precipitated using centrifugation, the pellet was washed and finally dissolved in 50 μl of nuclease free water. Next, 5 μl of the resulting solution was mixed with 5 μl of primer/probe mix (Table 1) and 10 μl of qScript XLT One-Step RT-qPCR ToughMix (Quantabio, USA) and was analyzed using CFX96 Touch real-time PCR detection system (Bio-Rad, USA). The PCR program was performed as follows: 15 min at 50 °C for reverse transcription reaction followed by 5 min at 95°C, then, 50 cycles, each comprising 20 s at 95 °C, 20 s at 58 °C, and 30 s at 72 °C. For each sample amplification of two different regions (N2 and N3) of the SARS-CoV-2 nucleocapsid (N) gene was analyzed in duplicates. For SARS-CoV-2 RNA copy number estimation serial 10-fold dilutions of standard samples with known concentration were used for standard curve generation and linear relationship between Ct values and amplicon copy number was observed for both PCR systems with lower limit of detection being 1000 copies in PCR reaction (Figure 1). Synthetic DNA fragments containing N2 and N3 regions and viral genomic RNA (kindly provided by Inna Dolzhikova, N.F. Gamaleya Research Center for Epidemiology and Microbiology, Moscow, Russia) were used for standard samples generation. SARS-CoV-2 RNA copy number in samples was calculated as the mean of four measurements (two values for N2 and two for N3) and converted to the total volume of the transport media thus reflecting the total viral RNA quantity in swab. Each PCR plate contained negative control samples without matrix RNA/DNA which served as an indicator of the absence of contamination.

**Table 1.**
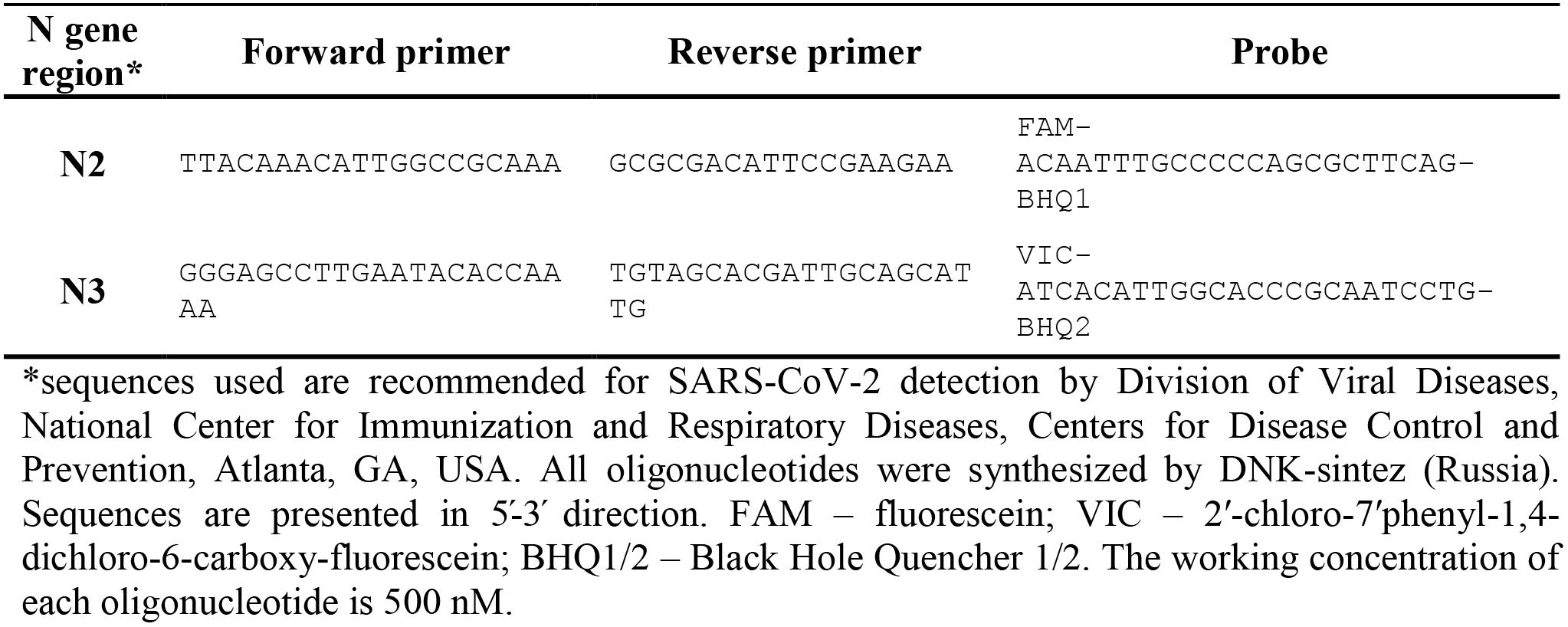
Oligonucleotides used for quantitative real-time PCR analysis.

**Figure 1.**
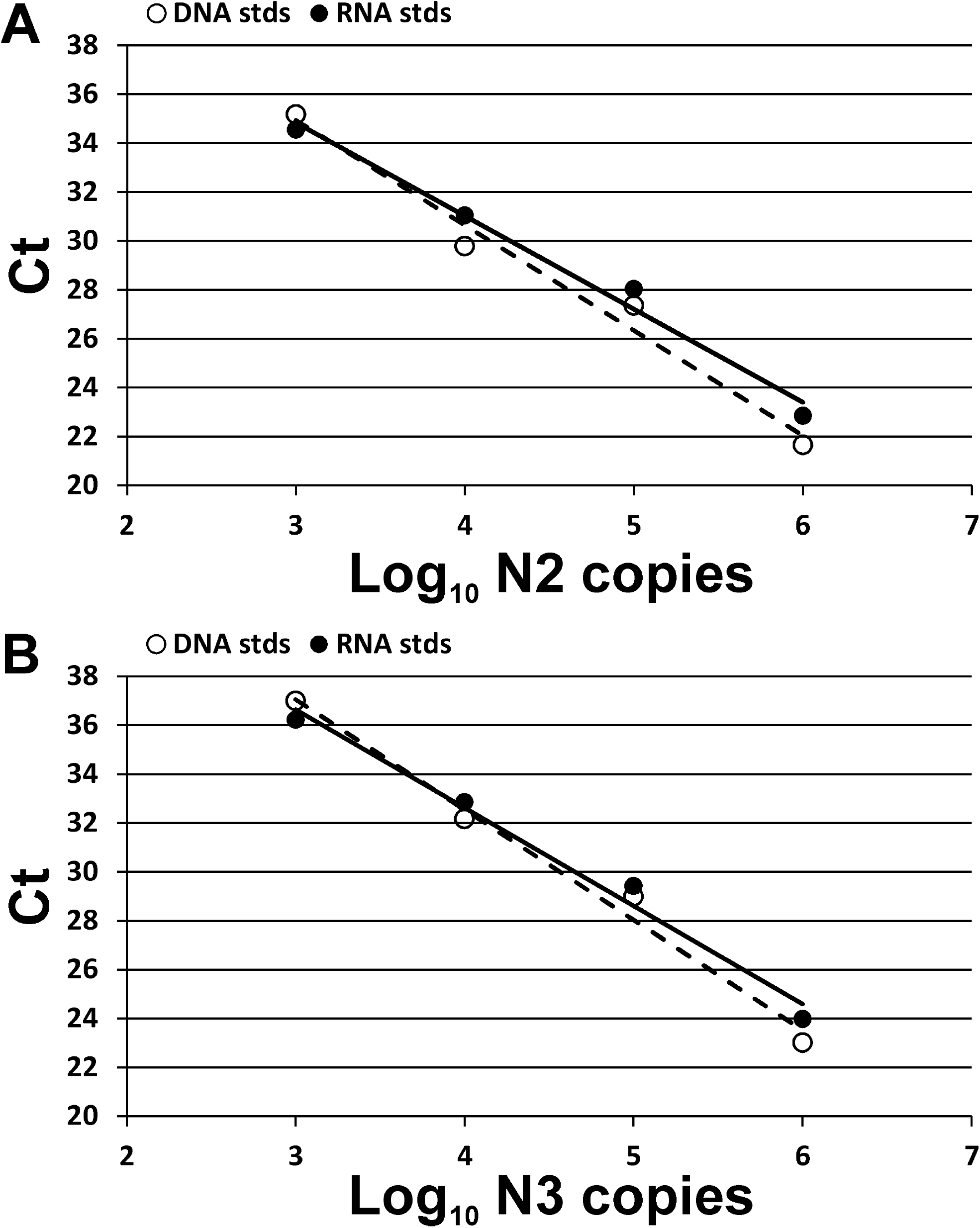
Standard curves used for calculating the SARS-CoV-2 RNA absolute number in swabs. Two PCR systems for detection of N2 (A) or N3 (B) regions of SARS-CoV-2 nucleocapsid (N) gene were used. The curves were generated using standard samples with either synthetic DNA fragments (white dots, dashed lines) or viral genomic RNA (black dots, solid lines). Both standards demonstrate similar results.

Statistical analysis was performed with Python 3 programming language with numpy, scipy and pandas packages. The Fisher exact test (two-tailed) was used for comparing qualitative parameters between independent groups of patients; the significance level α for p-values was set to 0.05. The Mann–Whitney U test (two-sided, with continuity correction) was used for comparing distributions of quantitative parameters between independent groups of patients. In order to control type I error, false discovery rate q-values was calculated using Benjamin–Hochberg (BH) procedure, and threshold of 0.05 was set to keep positive false discovery rate below 5%.

## Results and Discussion

In the current study 45 patients with COVID-19 in mild condition were analyzed for SARS-CoV-2 RNA in nasopharynx. On the initial day of the study (“day 0”) peripheral blood was collected and complete blood count as well as biochemical panel were analyzed. Nasopharynx swabs were taken on day 0, 3, and 8. Among patients included into the study 33 were receiving HCQ while 12 patients were receiving no antiviral pharmacological therapy. The age ranges of patients in control and experimental groups were comparable, but the groups differed in sex ratio (Table 2). Nevertheless, the key complete blood parameters and biochemical characteristics were indistinguishable between the groups except for red blood cell distribution width (RDW, p < 0.05) and total cholesterol (TH, p < 0.05); however, all the parameters including RDW and TH were in the normal range (Table 2).

**Table 2.** General characteristics of experimental groups.

| Parameter | Control (n = 12) | HCQ (n = 33) | p-value |
| --- | --- | --- | --- |
| Males/Females, % | 17/83 | 60/40 | NA |
| Age, years | 53.0 [44.8;60.8] | 49.0 [38.0;57.0] | 0.23 |
| Hemoglobin, g/L | 124.0 [123.0;132.0] | 141.0 [131.0;151.0] | 0.079 |
| Erythrocytes, 10<sup>12</sup>/L | 4.4 [4.3;4.6] | 4.8 [4.4;5.0] | 0.16 |
| Mean corpuscular volume, fL | 86.0 [84.0;88.0] | 89.0 [87.0;91.0] | 0.086 |
| Mean corpuscular hemoglobin, pg | 29.6 [27.8;29.8] | 30.0 [29.6;30.9] | 0.087 |
| Red blood cell distribution width, % | 14.1 [13.3;14.1] | 12.6 [12.4;13.1] | **0.032** |
| Mean corpuscular hemoglobin concentration, g/dL | 33.8 [33.2;34.0] | 34 [33.6;34.3] | 0.23 |
| Platelets, 10<sup>9</sup>/L | 239.0 [223.0;309.0] | 231 [173.0;268.0] | 0.15 |
| Leukocytes, 10<sup>9</sup>/L | 6.9 [5.1;8.5] | 5.1 [4.4;6.3] | 0.079 |
| Neutrophils, % | 61.4 [59.0;65.7] | 52.3 [46.5;59.9] | 0.087 |
| Lymphocytes, % | 27.7 [26.0;32.0] | 39.9 [30.9;44.3] | 0.087 |
| Monocytes, % | 7.0 [3.5;8.6] | 6 [4.9;6.6] | 0.31 |
| Eosinophils, % | 2.0 [1.5;2.3] | 1.4 [1.1;1.8] | 0.15 |
| Basophils, % | 0.5 [0.5;0.6] | 0.5 [0.4;0.6] | 0.34 |
| Creatinine, µM | 91.0 [80.0;100.0] | 92 [84.0;111.0] | 0.27 |
| Total cholesterol, mM | 4.6 [4.2;5.7] | 3.5 [3.1;4.0] | **0.048** |
| Triglycerides, mM | 1.5 [1.0;1.8] | 1.3 [0.9;1.8] | 0.45 |
| HDLp, mM | 1.1 [0.9;1.2] | 0.8 [0.7;1.1] | 0.079 |
| LDLP, mM | 3.2 [2.4;3.8] | 1.9 [1.4;2.2] | 0.079 |
| Total bilirubin, µM | 7.5 [5.4;9.7] | 10.3 [8.4;12.3] | 0.11 |
| Direct bilirubin, µM | 1.7 [1.2;1.8] | 1.8 [1.4;2.2] | 0.26 |
| Indirect bilirubin, µM | 5.8 [4.2;7.4] | 9 [6.7;10.6] | 0.087 |
| Alanine aminotransferase, IU/L | 21.0 [17.0;24.0] | 27 [21.0;38.0] | 0.15 |
| Aspartate aminotransferase, IU/L | 17.0 [15.0;19.0] | 16 [11.0;22.0] | 0.45 |
| Glucose, mM | 5.6 [5.0;7.5] | 5.8 [5.3;6.3] | 0.38 |
| C-reactive protein, mg/L | 0.1 [0.1;5.6] | 3.3 [0.1;17.4] | 0.25 |

HCQ - hydroxychloroquine. Values presented as median [quartile 2;quartile 3]. Differences between groups were quantified using Mann–Whitney U test with Benjamini-Hochberg correction. Significant differences ( $p < 0.05$ ) are marked in bold. NA – p-level cannot be calculated.

We found that total viral RNA quantity in nasopharynx swabs of all patients included into the study on day 0 was ranging from 0 (one patient) to 10^9^ copies, with median and interquartile range 150 000 [25 000 – 1 000 000]. RNA copies distribution had a maximum shifted towards 10 000 copies and prolonged right shoulder (Figure 2A). Measurement of viral RNA in the course of the disease revealed that on day 3 for most of the patients (68.9 %, 31 patients out of 45) RNA copies decreased, mostly more than 4 times (53.3 %, 24 patients out of 45). Meanwhile, the increase in RNA load was observed in 22.2% (10 of 45) patients, in some of them (6.7%, 3 of 45) more than 4 times. Furthermore, we found that higher RNA quantity on day 0 was correlated with greater fold change between days 0 and 3 (Figure 2B). This, together with the elimination of a significant proportion of patients with high viral load due to hospitalization, resulted in a significant narrowing of the RNA copies range at the day 3 and its “compression” around the value of ~10 000. We found even more narrow RNA load distribution on day 8 of the study with 22.2% (10 of 45) of patients having negative swabs (Figure 2A). These results may indicate that RNA copy number in the order of 10^4^ is associated with the recovery stage of infection. In this respect the results of our study are in agreement with previously published data where it was found that throat and nasal swabs of 2 11 infected individuals are characterized by RNA copy number varying from 10^2^ to 10^11^ [4-6].

**Figure 2.**
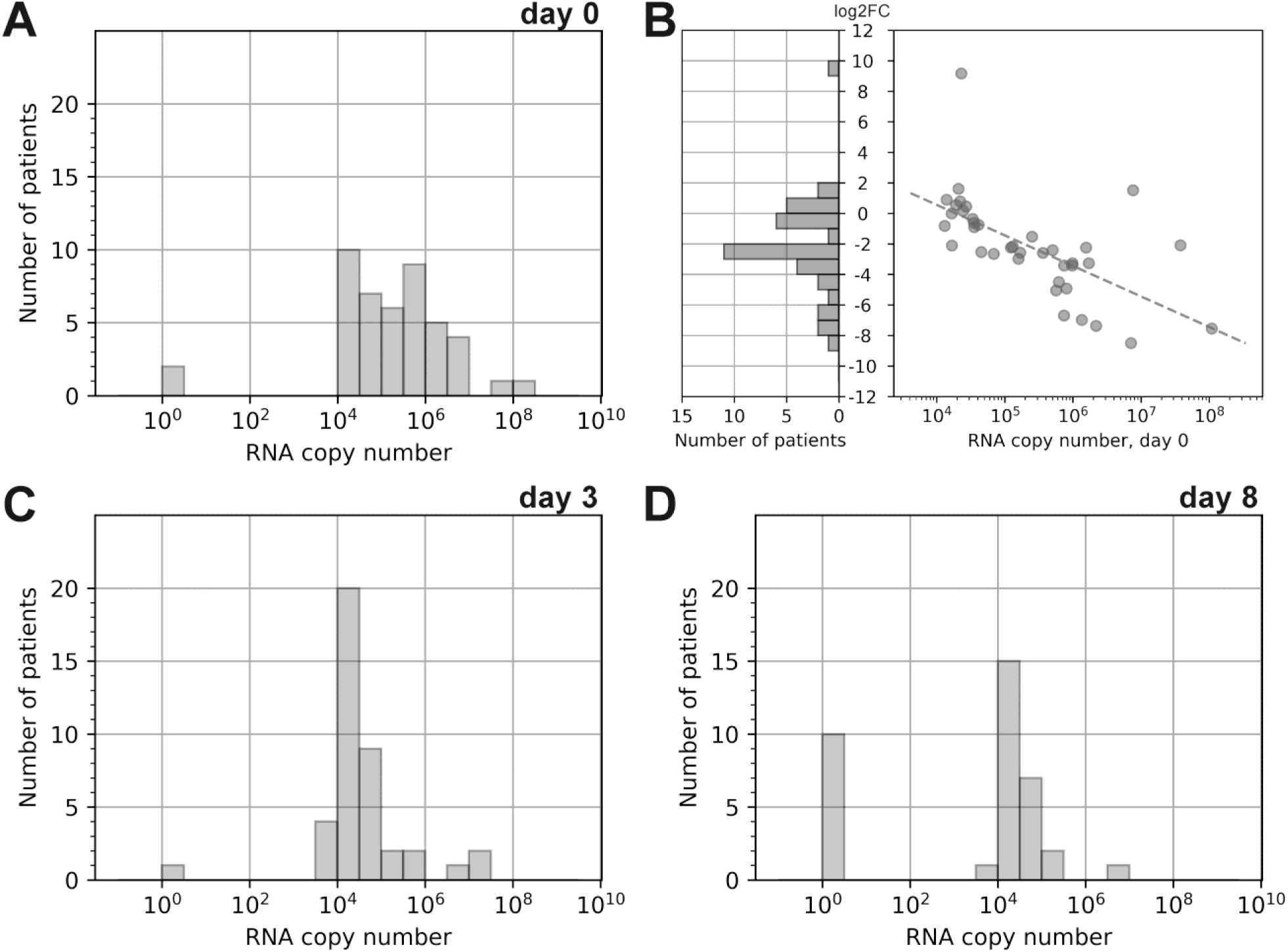
SARS-CoV-2 RNA quantity change dynamics. The distribution of viral RNA load in nasopharynx of patients at day 0 (A), day 3 (C) and day 8 (D). To use the logarithmic scale the exact zero was replaced with one (10^0^). Distribution of log2 transformed fold change of RNA copy number between swabs on day 0 and 3 (B Left). Dependence between log2 transformed of RNA copy number fold change and RNA copy number on day 0; dashed line shows linear approximation of the trend (Spearman correlation, r = −0.7, p-value < 0.00001) (B Right). Only patients that have non-zero values of RNA copy number both on days 0 and 3 and were not hospitalized between days 0 and 3 are shown.

Although direct comparison between the results of these studies and our work is complicated due to different experimental procedures, similar trends were observed, e.g., in all studies recovering patients demonstrated considerably lower RNA copy number compared with productive stage of infection. Additionally, the results of the current study indicate that recovering patients may produce viral RNA even after 18 days after the onset of symptoms. Similar results were demonstrated by Zhou F et al. [7] for patients with COVID-19 in Wuhan, China – in the study median duration of viral shedding was found to be 20 days (IQR 17-24), with the longest observed duration of viral shedding being 37 days.

When viral RNA dynamics in individual groups were compared, there were no statistically significant correlations between decrease/increase in RNA copy number and the HCQ administration – in each time point the RNA load in swabs was comparable between the groups (Table 3). Moreover, on day 8 (end of the observation period in the current study) 33.3% (4 of 12) and 18% (6 of 33) of patients were characterized by negative swabs in control and HCQ receiving groups, respectively. Taken together these results demonstrate that HCQ therapy neither shortened the viral shedding period nor reduced the virus RNA load.

**Table 3.** Comparison of viral RNA load at different time points between groups.

|  | Control group |  | HCQ group |  | Mann-Whitney U Test |  |
| --- | --- | --- | --- | --- | --- | --- |
|  | n | median<br>IQR[Q2;Q3] | n | median<br>IQR[Q2;Q3] | p-value unadj | q-value |
| Day 0, total RNA<br>copies in swab | 12 | 447 847<br>[23 821;1 030 282] | 33 | 130 183<br>[33 812;982 481] | 0.38 | 0.45 |
| Day 3, total RNA<br>copies in swab | 12 | 27 638.5<br>[20 138;45 794] | 29 | 27 715.0<br>[17 204;87 433] | 0.45 | 0.45 |
| Day 8, total RNA<br>copies in swab | 10 | 19 001.5<br>[0;28 313] | 26 | 23 880.5<br>[8 094;36 859] | 0.18 | 0.45 |

HCQ – hydroxychloroquine; IQR – interquartile range; Q2 – quartile 2; Q3 – quartile 3. Differences between groups were quantified using Mann–Whitney U test without (p-value unadj) and with Benjamini-Hochberg correction (q-value).

Further analysis has shown no statistically significant correlations between maximal RNA quantity in swabs and patients’ age, sex, or blood parameters. However, we found significant relationship between maximal RNA quantity and patients’ deteriorating medical conditions, as defined in Methods. Patients with RNA load higher than 10^6^ copies significantly more frequently showed deterioration in their condition compared to those with RNA load below 10^6^ copies (p = 0.000066, Fisher exact test) (Table 4). Among hospitalized patients only one belonged to the control group while the rest (9 of 10) were receiving HCQ; however, this difference may be the result of a smaller size of the control group. Hospitalization cases also showed strong positive correlation with viral RNA load: patients with higher than 10^6^ viral RNA copies were hospitalized significantly more frequently compared to those with RNA load below 10^6^ copies (p = 0.0029, Fisher exact test) (Table 4). The age of patients and blood parameters were comparable between the groups of hospitalized/non-hospitalized. However, in agreement with recently published data [8] we found that patients with arterial hypertension were hospitalized significantly more frequently compared to patients without hypertension (p = 0.009, Fisher exact test) (Table 4), though there was no correlation between this parameter and viral RNA load.

**Table 4.** Significant correlations between RNA quantity and patients’ clinical data.

|  | **no deterioration** | **deterioration** | **p-value** |
| --- | --- | --- | --- |
| **RNA copy number $\leq 10^6$** | 26 | 6 | 0.000066 |
| **RNA copy number $> 10^6$** | 2 | 11 |  |
|  | **non-hospitalized** | **hospitalized** | **p-value** |
| **RNA copy number $\leq 10^6$** | 29 | 3 | 0.0029 |
| **RNA copy number $> 10^6$** | 6 | 7 |  |
|  | **non-hospitalized** | **hospitalized** | **p-value** |
| **no arterial hypertension** | 27 | 8 | 0.0091 |
| **arterial hypertension** | 3 | 7 |  |

p-values were determined using Fisher's exact test.

## Conclusions

We found that there is a statistically significant positive correlation between SARS-CoV-2 RNA quantity in nasopharynx and deterioration of patients’ medical conditions leading to hospitalization. Hydroxychloroquine administration has no statistically significant effect on SARS-CoV-2 RNA in nasopharynx of patients with COVID-19. In accordance with the recent Food and Drug Administration’s comment we confirm that the use of hydroxychloroquine for the infection treatment and prophylactics is doubtful. Although, our study has a significant limitation due the relatively small number of patients, our finding, together with the results of recently published works, indicates that quantitative PCR can be a perspective approach for monitoring of COVID-19 course and prediction of deterioration in patient’s condition.

## Data Availability

All data generated and analysed during the study are included in the article. Any additional information is available from the corresponding author on reasonable request.

## Acknowledgments

We thank Dr. Inna Dolzhikova (N.F. Gamaleya Research Center for Epidemiology and Microbiology, Moscow, Russia) for providing with SARS-CoV-2 genomic RNA used for the calculation of absolute number of viral RNA in samples. This study was funded by the Russian Science Foundation grant, agreement #18-15-00420.

## References

1. Yao X, et al. In vitro antiviral activity and projection of optimized dosing design of hydroxychloroquine for the treatment of severe acute respiratory syndrome coronavirus 2 (sars-cov-2). Clinical infectious diseases: an official publication of the Infectious Diseases Society of America. 2020;71:732–739

2. Liu J, et al. Hydroxychloroquine, a less toxic derivative of chloroquine, is effective in inhibiting sars-cov-2 infection in vitro. Cell discovery. 2020;6:16

3. Hashem AM, et al. Therapeutic use of chloroquine and hydroxychloroquine in covid-19 and other viral infections: A narrative review. Travel medicine and infectious disease. 2020;35:101735

4. Pan Y, Zhang D, Yang P, Poon LLM, Wang Q. Viral load of sars-cov-2 in clinical samples. The Lancet Infectious Diseases. 2020;20:411–412

5. Zou L, et al. Sars-cov-2 viral load in upper respiratory specimens of infected patients. New England Journal of Medicine. 2020;382:1177–1179

6. Yu F, et al. Quantitative detection and viral load analysis of sars-cov-2 in infected patients. Clinical infectious diseases: an official publication of the Infectious Diseases Society of America. 2020;71:793–798

7. Zhou F, et al. Clinical course and risk factors for mortality of adult inpatients with covid-19 in wuhan, china: A retrospective cohort study. The Lancet. 2020;395:1054–1062

8. Huang S, et al. Covid-19 patients with hypertension have more severe disease: A multicenter retrospective observational study. Hypertension Research. 2020;43:824–831

